# 2D Transfer Learning for ECG Classification using Continuous Wavelet Transform

**DOI:** 10.1101/2024.07.11.24310258

**Authors:** Wei Zhang

## Abstract

Advanced deep neural networks, when trained on extensive datasets, can outperform cardiologists in diagnosing cardiac arrhythmias. However, the availability of large-scale training data is often impractical. This study explores the use of transfer learning to identify and classify three ECG patterns. It applies knowledge gained from 2D image classification tasks to the domain of 1D time-series ECG signal classification. The research leverages various deep learning models to classify continuous wavelet transform (2D representations) of ECG signals. The effectiveness of these transferred deep learning models in classifying ECG time-series data is then evaluated.

## 1 Introduction

Cardiovascular diseases (CVDs) are the leading cause of mortality globally, early CVD diagnosis is crucial for preventing premature deaths. While standard diagnostic techniques such as angiography, magnetic resonance imaging (MRI), nuclear imaging, and echocardiography are costly, electrocardiogram (ECG) offers an inexpensive and widely accessible alternative with the potential to save millions of lives. The labor-intensive and challenging nature of visual ECG analysis has necessitated the development of Computer-Aided Diagnosis (CAD) systems.

Recent advancements in deep learning techniques have shown significant potential in ECG signal classification. Recurrent Neural Networks (RNN), One Dimensional Convolutional Neural Network (1DCNN) [1, 2], and Long Short Term Memory (LSTM) Network [3, 4] have demonstrated remarkable performance in this field. Despite the ability of deep learning models to outperform cardiologists, they require extensive datasets for robust training. One potential approach to address this data scarcity is the generation of synthetic training data. However, models trained on synthetic data may struggle to predict specific trends within the database [5]. Transfer learning emerges as another promising approach to overcome this limitation.

Transfer learning has significant potential to enhance CNN-based classifiers’ performance when large training datasets are unavailable. Recent applications of transfer learning to ECG signals include epilepsy diagnosis [6], fall detection [7], and arrhythmia classification [8]. [9] conducted a comprehensive validation showing that transfer learning was effective for small ECG datasets. However, a analysis based on 2D architecture depth for time-series signal classification is not yet available. This research aims to address this gap.

In this work, we compare seven established deep learning architectures of varying depths, ranging from 41 to 708 layers. The study utilizes scalograms instead of other transformations such as Gramian Angular Summation Field (GASF), as scalograms retain more fine-grain features. We extensively analyze the transferability of ImageNet pre-trained models to time-series ECG signals, providing a novel contribution to the field of ECG signal analysis and classification. The primary contribution of this work is to assess the transferability of various general image classification architectures for classifying time-series signals, specifically ECG signals.

## 2 Datasets

This research utilizes ECG data collected from three groups of individuals, comprising a total of 162 ECG recordings from the MIT-BIH Normal Sinus Rhythm Database [10, 11]. This database contains recordings from 47 subjects with arrhythmia, studied by the BIH Arrhythmia Laboratory at Boston’s Beth Israel Hospital.

This diverse dataset allows for a comprehensive analysis of various cardiac conditions using deep learning techniques.

## 3 Methods

### 3.1 Continuous Wavelet Transform

The Continuous Wavelet Transform (CWT) is a powerful tool for analyzing localized variations of power within a time series. Unlike the Fourier Transform, which provides a global representation of the signal’s frequency content, the CWT offers a time-frequency representation, making it ideal for analyzing non-stationary signals.

The CWT of a signal *x*(*t*) is defined as the convolution of *x*(*t*) with a scaled and translated version of a wavelet function *ψ*(*t*):

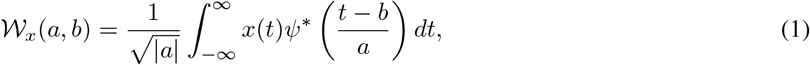

where *a* is the scaling parameter, *b* is the translation parameter, and *ψ*^*^(*t*) denotes the complex conjugate of the mother wavelet *ψ*(*t*). The parameters *a* and *b* allow the wavelet to be dilated and shifted, providing a detailed time-frequency analysis.

The choice of the mother wavelet *ψ*(*t*) is crucial in CWT analysis, as it determines the features of the signal that will be captured. Commonly used wavelets include the Morlet wavelet, the Mexican Hat wavelet, and the Haar wavelet. Each wavelet has its own characteristics and is suited to different types of signal features.

For instance, the Morlet wavelet, which is a complex exponential modulated by a Gaussian, is particularly useful for detecting oscillatory behavior in the signal:

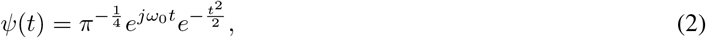

where *ω*_0_ is the central frequency.

### 3.2 Transfer Learning

Training a deep CNN prediction model from scratch requires substantial training data and computational power. In many applications, an adequate quantity of training data is unavailable. In such scenarios, reusing available CNNs trained on large datasets for conceptually similar tasks proves beneficial.

The principle of transfer learning is based on the idea that knowledge learned from patterns in one domain may apply to other domains. Transfer learning utilizes a pre-trained deep neural network as leverage for automated feature extraction. The convolution layers within this model contain features learned during initial training and possess knowledge about the patterns in the original dataset. These feature representations can serve as feature extractors for a new dataset [12].

The extracted features from the mid-layers of a deep CNN are robust enough to capture hand-engineered features and are ideal for feature extraction [13]. The transfer learning technique has great potential for automation in various domains such as sound signals [14], aircraft design [15, 16], and medical application development cycles [17, 18].

Figure 1 illustrates the transfer learning procedure used in this work.

**Figure 1:**
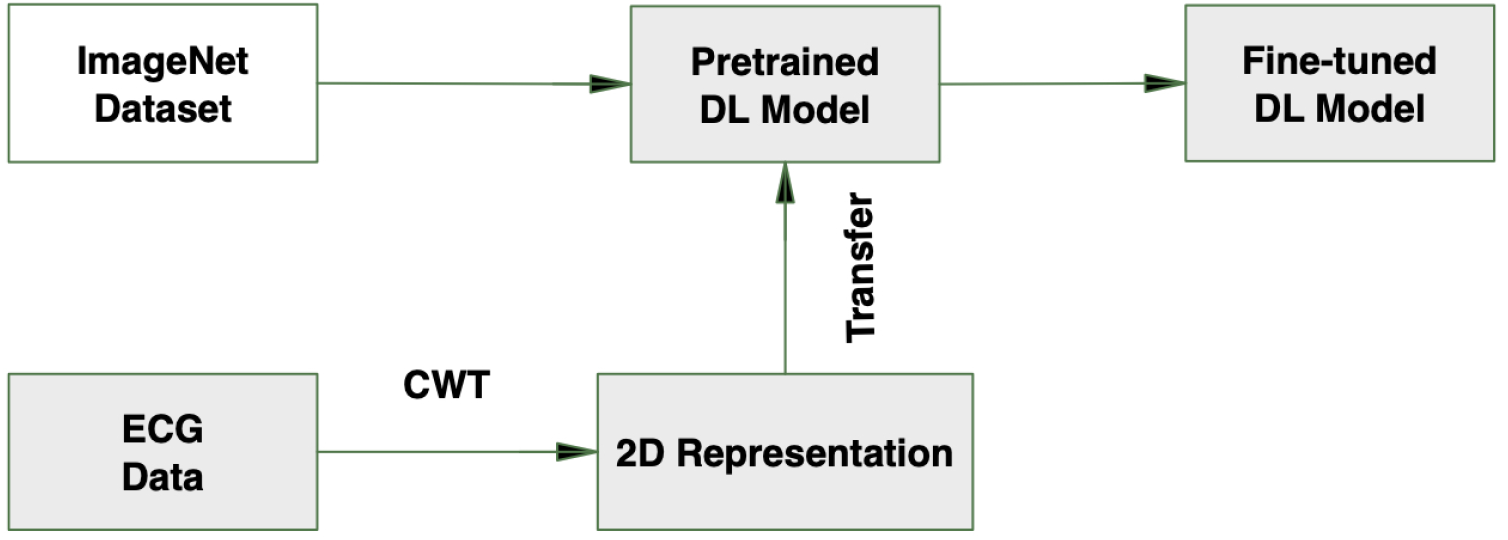
Transfer Learning procedure.

### 3.3 Experiment

The feature extraction process occurs layer by layer, beginning with the input, progressing through hidden layers, and culminating at the final classification output layer. Our approach involves modifying the pre-trained models by replacing their last three layers with a fully-connected layer, an activation layer (ReLU), and a softmax layer. The input image resolution is adjusted to meet the specific requirements of each model.

The learning process is initiated with a higher learning rate to accelerate convergence. The fully-connected layer elongates the feature map into a vectorized form, which is then followed by a multi-class activation function suitable for classification problems. We employ categorical cross-entropy as the cost function to optimize the pre-trained models.

For hyperparameter selection, we have utilized a heuristic approach. Among the tested optimizers, RMSprop demonstrated the highest performance. This methodology allows us to leverage the power of pre-trained models while adapting them to our specific ECG classification task.

## 4 Results & Discussions

We conducted a comprehensive analysis using various state-of-the-art pre-trained architectures of different depths. Conventionally, the performance of a deep network improves with increasing architecture depth, as each layer extracts essential features that serve as input to the next level.

Our simulations revealed that increasing depth contributed to accuracy gains up to a certain point, after which performance degraded. The ECG classification task achieved the highest response with ResNet-50, obtaining an accuracy of 94.12%. This finding motivates the development of a transfer learning approach with reduced complexity.

The proposed approach aims to overcome the data scarcity problem in medical diagnostics through transfer learning. Table 2 presents the performance of various architectures. The superior performance of ResNet-50 and GoogLeNet can be attributed to their inherent domain-independent generalization ability. Despite DenseNet’s [19] general efficiency compared to ResNet, its poor performance in this context results from its fundamental difference in using concatenation operations instead of addition. Consequently, DenseNet is not a suitable candidate for time-series analysis in this context.

**Table 1:**
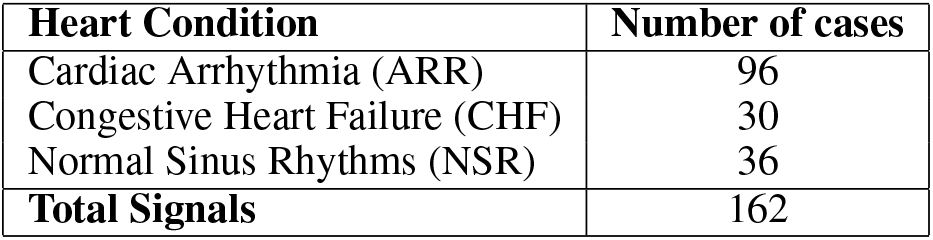
Description of the Dataset.

| Heart Condition | Number of cases |
| --- | --- |
| Cardiac Arrhythmia (ARR) | 96 |
| Congestive Heart Failure (CHF) | 30 |
| Normal Sinus Rhythms (NSR) | 36 |
| **Total Signals** | **162** |

**Table 2:** ECG Classification Results Using Transfer Learning.

| Model | Accuracy |
| --- | --- |
| ResNet-18 [20] | 88.24% |
| EfficientNet-B0 [21] | 85.93% |
| GoogLeNet [22] | 91.03% |
| VGG 16 [23] | 89.62% |
| ResNet-50 [20] | 92.72% |
| DenseNet-201 [19] | 83.04% |

These findings underscore the importance of carefully selecting pre-trained architectures for transfer learning in medical image classification tasks, particularly when dealing with time-series data like ECG signals.

## 5 Conclusion

The primary advantage of transfer learning is its ability to rapidly adapt a deep learning model with proven performance to a new domain, eliminating the need to develop a new model from scratch. This approach significantly reduces the computational cost associated with determining complex layer parameters and lengthy validation processes. The similarity in accuracy among the latest deep learning algorithms suggests a comparable low-level feature extraction capability across these significantly different architectures. While the achieved accuracy is commendable, this work underscores the importance of high-level abstract features. This comprehensive comparison of deep-transfer between various architectures provides valuable insights for informed architecture selection in time-series data classification tasks. The implications of this research extend beyond ECG signal analysis and can be applied to the detection of other chronic cardiovascular diseases, including cardiomyopathy, coronary heart disease, and structural abnormalities. Future research directions could focus on enhancing model response through fine-tuning, potentially leading to more accurate and efficient diagnostic tools.

## Data Availability

All data produced in the present work are contained in the manuscript

